# When Data Meets Practice: A Qualitative Study of Clinician Perspectives on Streaming Data in Mental Health

**DOI:** 10.64898/2026.04.23.26351640

**Authors:** Julie Tian, Viktoriia Kurkova, Yiqing Wu, Medard Adu, Jake Hayward, Andrew J. Greenshaw, Bo Cao

## Abstract

Patient-generated streaming data from wearable and digital technologies is increasingly promoted as a means of supporting mental health monitoring and clinical decision-making. While patient acceptance of these technologies has been reported, clinician perspectives remain underexplored despite their central role in determining whether streaming data are meaningfully integrated into routine care. This study explored clinicians’ experiences, as well as perceived facilitators and barriers, related to integrating patient-generated streaming data into routine mental health practice.

A qualitative, exploratory interview study was conducted to examine clinicians’ experiences and perspectives on integrating patient-generated streaming data into mental health care. Semi-structured interviews were conducted with 33 clinicians, including family physicians (n=11), psychiatrists (n=12), and psychologists (n=10). Data were analyzed using reflexive thematic analysis guided by Braun and Clarke’s six-step approach.

Six themes were identified. Clinicians described variable use of digital and streaming technologies, ranging from routine engagement to deliberate non-use. Streaming data were viewed as clinically valuable when they provided longitudinal and objective insights, identified physiological and behavioural pattern changes, and supported patient engagement. However, clinicians emphasized that clinical usefulness was contingent on interpretability, contextual information, and relevance to decision-making. Major barriers included poor integration with electronic medical records, time constraints, data volume, limited organizational support, and uncertainty regarding data reliability and validity. Clinicians also expressed persistent concerns about privacy, governance, and regulatory oversight, highlighting the need for clear safeguards and accountability structures.

Clinicians view patient-generated streaming data as a promising adjunct to mental health care, particularly for capturing longitudinal change between visits. However, meaningful clinical integration remains constrained by usability, workflow, organizational, and regulatory challenges, as well as limited confidence in data interpretation. Addressing these barriers through improved system integration, interpretive support, validation, and governance will be essential for translating the potential of streaming data into routine clinical practice.

**Author Summary:** Mental health symptoms can change between appointments yet care often depends on periodic visits and patient recall. Devices such as smartwatches and other digital tools can continuously collect information, from mood and sleep to activity and related measures, offering a possible way to support care outside the clinic. While patients are often seen as the main users of these tools, clinicians play a central role in deciding whether such technology is implemented in care. This study interviewed 33 mental health clinicians, including family physicians, psychiatrists, and psychologists, about their views on using patient-generated streaming data in routine care. Clinicians saw promise in these data as they help track changes over time, support discussions with patients, and provide additional insight between visits. However, they also described important barriers, including managing large amounts of data, limited integration with health record systems, uncertainty about data quality, and concerns about privacy and regulation. These findings suggest that successful implementation of streaming data in mental health care will depend on designing systems that are clinically relevant, easy to interpret, and supported by appropriate safeguards and infrastructure.

## Introduction

Mental disorders are common conditions, yet they remain a persistent contributor to population-level morbidity and functional impairment. Globally, mental disorders continue to rank among the leading causes of non-fatal disease burden, with the Global Burden of Disease 2019 analysis showing no evidence of a sustained reduction in burden since 1990 [1]. In Canada, mental disorders are similarly prominent in national burden estimates, with mental disorders being the second largest source of all-age years lived with disability, closely following musculoskeletal disorders [2]. At the same time, mental health systems continue to face structural constraints that limit timely access and continuity, contributing to ongoing gaps between care needs and care received. In 2018, 17.8% of Canadians reported needing mental health services, with 22.4% of those having their needs partially met and 21.4% fully unmet [3]. These gaps are consistent with persistent access barriers, including long waits for specialist services, financial barriers, a lack of professionals in the area, and difficulties finding a specialist [4,5].

In this context, digital health technologies have been advanced as one strategy to extend mental health support. The World Health Organization defines digital health as the use of information and communication technologies, such as mobile phones and health applications, wearable devices, and telemedicine, to deliver health information and services [6]. The expansion of digital health has brought increased focus to continuous, or streaming, data as a tool for generating objective measures of daily function and monitoring mental health. Streaming data describes real-time information that is continuously generated by connected digital devices, making it possible to repeatedly track physiological and behavioural signals across time [7–10]. Wearable devices are a key source of streaming data and commonly generate multiple metrics across domains, including sleep patterns, physical activity, heart rate, blood pressure, blood glucose, and other physiological indicators [11–14].

Current mental healthcare relies heavily on episodic appointments and retrospective self-report, approaches that are vulnerable to recall bias and often miss meaningful changes that occur between visits [15]. Illness management often requires more detailed monitoring capable of capturing early deviations that precede clinical deterioration; streaming data helps address this limitation by enabling continuous, real-time monitoring of physiological and behavioural patterns, making it possible to detect subtle deviations from an individual’s baseline before symptoms escalate. This is particularly relevant in mental illness, where early warning signs frequently precede relapse, and strategies that support early identification and response have been proposed as a way to prevent recurrence and reduce hospitalization [16]. In mood disorders, longitudinal wearable data are increasingly being used to detect patterns linked to episode onset, including personalized machine learning models using sleep and circadian rhythm features to predict future mood episodes [17] and findings that day-to-day variability in objectively measured activity may signal early transitions to depressive symptoms in bipolar disorder before changes in sleep or self-reported mood appear [18]. Thus, patient-generated streaming data can support clinical monitoring and decision-making.

To date, research on streaming data and wearable technologies in mental health has largely focused on patient outcomes, feasibility, and acceptability, with studies showing that patients are generally receptive to using wearables and passive monitoring tools, particularly when these technologies are perceived as low burden, supportive of self-awareness, and aligned with symptom management or relapse prevention [19–21]. Far less attention has been paid to clinicians’ perspectives, beyond broad surveys or technology-specific evaluations. Yet, the integration of streaming data into routine care ultimately depends on clinicians, who decide whether patient-generated data are discussed during visits, how they are interpreted in clinical context, and whether they inform care decisions [22,23].

If poorly integrated, streaming data systems can contribute to clinician burnout, technostress, and workflow strain, especially when large volumes of data must be interpreted under time constraints [24–26]. Broader implementation research shows that barriers to digital mental health technologies are multi-level, including training gaps, infrastructure limitations, and unresolved privacy and security concerns [27]. Adoption is further shaped by patients’ perceptions of usefulness and usability, as well as clinicians’ and policymakers’ views on data reliability, workflow impact, and professional responsibility [28–30]. Tools developed without meaningful end-user input often misalign with clinical practice and face low uptake or discontinuation, whereas sustained stakeholder engagement improves relevance, usability, and trust, increasing the likelihood of successful implementation [31–33]. This underscores the need to directly examine clinicians’ perspectives, as their experiences and judgments are central to whether streaming data becomes a meaningful part of care.

Taken together, existing literature positions clinicians as key clinical decision-makers and implementation stakeholders in if and how patient-generated streaming data is incorporated into mental health care [22,23,34,35]. Clinicians determine if these data are reviewed during encounters, how they are interpreted alongside clinical history and patient report, and whether they meaningfully inform risk assessment or treatment planning. Yet clinician perspectives remain insufficiently understood; more exploration is needed regarding what they consider clinically valuable, how data review can fit into real-world workflows, and what safeguards are needed to support responsible use. This qualitative interview study examines clinicians’ experiences with and expectations for streaming data in mental health care, focusing on perceived value, usability, and workflow fit, feasibility and system barriers, data quality, and privacy/regulation considerations. Addressing this gap is essential, as clinician engagement ultimately determines whether patient-generated streaming data can move beyond research and pilot implementations to inform real-world mental health care.

## Methods

This qualitative, exploratory study examined clinicians’ perspectives on integrating patient-generated streaming data (e.g., wearable-derived sleep, activity, and physiological indicators) into routine clinical mental health care. Participants were invited to complete a 30-minute semi-structured interview via Zoom to explore their experiences, perceived barriers and facilitators, and practical use cases shaping the feasibility of adopting streaming data in day-to-day clinical workflows. Ethics approval was obtained from the University of Alberta’s Health Research Ethics Board (Pro00155136) prior to the commencement of research activities. E-consent was obtained from all participants prior to study participation.

### Participant Inclusion and Exclusion Criteria

Eligible participants included currently or previously licensed family physicians, psychiatrists, clinical psychologists, and residents/trainees in these specialties who currently provide, or previously provided, direct patient care in clinical settings serving individuals with mental health concerns. Participants were required to be able to provide informed consent in English and complete a virtual interview using Zoom. Clinicians from other medical specialties, individuals unable to provide informed consent, non-English speakers, and those unable to commit to the interview duration were excluded. Clinicians were recruited through professional networks, relevant associations, and introductions facilitated by the research team and supervisory committee. Interested participants completed electronic informed consent via a secure REDCap e-consent process before any study procedures commenced.

### Data Collection

Semi-structured virtual interviews were conducted via Zoom by JT, throughout October 2025, using a discussion guide that explored clinicians’ experiences and perceptions regarding streaming data, including perceived clinical utility, workflow integration, preferred data types and visualization, feasibility, privacy considerations, and data sharing. Participants were informed that they could decline to answer any question or stop the interview at any time without consequence. To support confidentiality, participants’ Zoom display names were changed to their assigned participant number prior to starting the recording of the session, and no patient-identifying information was solicited. All interviews were audio-recorded, transcribed verbatim, and de-identified by removing identifying details from transcripts. Interview recordings and transcripts were stored on encrypted, access-controlled University of Alberta servers.

### Data Analysis

Data were analyzed using reflexive thematic analysis guided by Braun and Clarke’s six-step approach (familiarization, coding, theme development, theme review, theme definition, and reporting) [36]. A research team member (JT) verified transcript accuracy and anonymization prior to analysis, after which transcripts were thematically analyzed through an iterative process. Using an inductive approach, JT and VK independently reviewed and coded the first transcript line-by-line to generate initial codes directly from the data. The research team (JT, VK, YW) met to compare interpretations, resolve discrepancies through discussion, and develop an initial coding framework, which included code definitions and application rules. This framework guided coding of subsequent transcripts and was iteratively refined as analysis progressed, with codes being created, collapsed, expanded, or redefined to reflect emerging concepts. Coding was conducted alongside analytic memoing and regular team meetings to support reflexivity and consistency in code application. Analysis proceeded until thematic saturation was reached, defined as the point at which no substantively new codes or themes emerged in later interviews. A third researcher (YW) independently reviewed the coded dataset and thematic structure and contributed to consolidating themes and resolving discrepancies. Final themes and subthemes were confirmed through iterative meetings and are reported with de-identified illustrative quotations. The Standard for Reporting Qualitative Research guided the reporting process [37].

## Results

A total of 36 clinicians consented to participate in the study. Of these, 33 completed an interview and were included in the analysis. Three clinicians did not complete the interview and were therefore excluded. The final analytic sample comprised 33 clinicians, including 11 family physicians, 10 psychologists, and 12 psychiatrists. Six themes were identified across the interviews (Figure 1). Clinicians first described their current use of digital and streaming technologies, which ranged from routine use of apps and wearable data in care to limited or no integration in practice. They then discussed the perceived clinical value of streaming data, highlighting where it may support clinical decision-making and where its usefulness is constrained. The third theme focused on usability, accessibility, and user support, emphasizing the importance of intuitive tools, patient and clinician burden, and the need for guidance to interpret data meaningfully. Participants also identified major challenges related to feasibility, system barriers, and workflow fit, including time constraints, integration into clinical processes, and restrictive infrastructure. Concerns regarding data quality, reliability, and validity emerged as central to whether streaming data could be trusted and acted upon in clinical care. Finally, clinicians highlighted privacy and regulation considerations, underscoring the need for clear governance, secure data handling, and regulatory frameworks to support responsible implementation.

**Figure 1.**
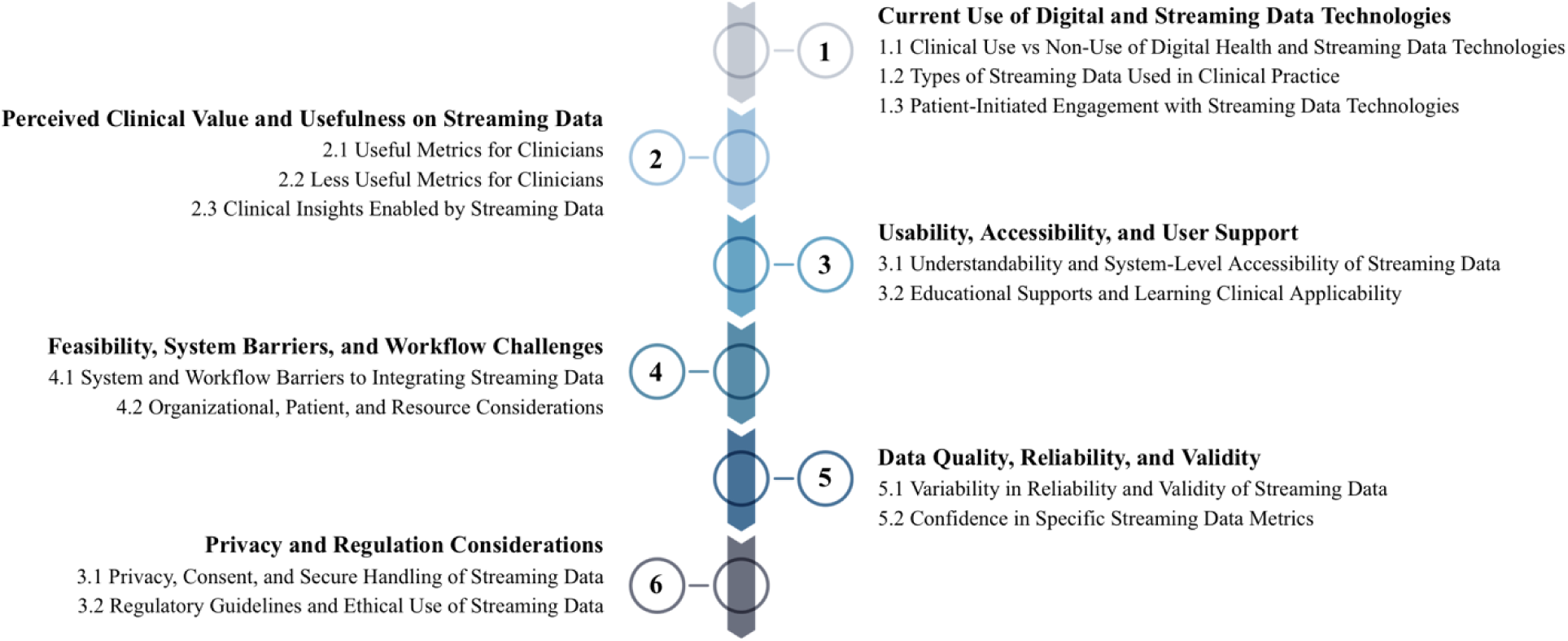
**Overview of Themes and Subthemes**. Thematic map of the six themes and its corresponding subthemes synthesized from clinician perspectives on streaming data in mental health care.

### Theme 1: Current Use of Digital and Streaming Data Technologies

Participants described varied engagement with digital and streaming data technologies in routine clinical care. While some clinicians reported actively recommending or reviewing select digital tools, others described minimal exposure or deliberate non-use. When streaming data were discussed, they were typically limited to a small subset of metrics, and the technology was most often introduced by clinicians rather than patients.

### Subtheme 1.1: Clinical Use vs. Non-Use of Digital Health and Streaming Data Technologies

Some clinicians described routinely recommending specific digital health applications or resources (n=14) to their mental health patients. Several clinicians use or recommend streaming data technologies (n=18) to support patients’ symptom monitoring or coping strategies between visits. These tools were viewed as accessible accompaniment that could enhance patient reflection without substantially altering clinical workflows.

*“We recommend using the fitness tracker to count their steps.” (Participant 4)*

*“I recommend various applications on the phone. So, this is things like MindShift, Headspace, Calm, stuff like that. Or as well as, an online website, like AnxietyCanada.com. There’s also a sleep hygiene website that I use that is called MySleepwell.ca.” (Participant 18)*

Some clinicians explicitly mentioned that streaming data was a regular part of their practice or have seen streaming data in practice (n=18) while other clinicians described limited and irregular engagement with patient-generated streaming data, noting that while they did not routinely recommend such tools, they would discuss them when relevant (n=9).

*“Patients will come and they’ll show us the logs and stuff that they do, and so that’s a regular part of the practice.” (Participant 23)*

*“People also use those wearables, like the Fitbit, and occasionally heart rate variability. Although I don’t recommend them actively, I will talk about them…I haven’t been doing that very regularly. And if I do recommend it, it’s fairly sporadic, it’s not with every single patient.” (Participant 28)*

In contrast, several clinicians reported not using or recommending digital health applications (n=9) or streaming data technologies (n=9) at all. Some of these participants cited lack of exposure to these technologies and uncertainty about their utility.

*“I haven’t made any recommendations around that, like smartwatches or anything like that. I haven’t directly seen patient-generated data used in clinical settings.” (Participant 14)*

*“I don’t actually use or recommend them, only because we haven’t had a lot of exposure to what to recommend and what’s out there.” (Participant 15)*

This variation underscored the absence of a shared framework or expectation regarding digital technology use across clinical settings.

### Subtheme 1.2: Types of Streaming Data Currently Used in Clinical Practice

When clinicians described engaging with streaming data technologies in clinical settings, their use was limited to a specific set of data metrics. Participants often referenced physiological data that was familiar, concrete, and aligned with existing clinical knowledge. Blood glucose data (n=8) was among the most frequently mentioned, along with heart rate data (n=8) recorded through smartwatches. Blood pressure (n=1) was the least frequently mentioned metric.

Clinicians noted the relevance of heart rate in the context of psychiatric medications and its perceived relationship to stress.

*“I would say the other one would be a smartwatch where they’re recording their heart rate. Again, psychiatric medications can affect the heart. And also, I think heart rate is a measure of how stressed someone can be.” (Participant 1)*

*“I have seen, definitely, the glucose monitoring. That would be the one that I’ve probably seen the most.” (Participant 12)*

In addition to physiological measures, some clinicians reported using sleep data (n=9) and self-reported mood tracking data (n=2) in therapeutic contexts. This data was described as being used actively within sessions to support reflection, evaluate change over time, and inform clinical priorities.

*“With some clients, if they consent to it, we use tracking of mood to use as evidence in therapy, to look at improvements, to challenge thought patterns versus evidence, and to address one-off incidences or trends in either direction, which helps change session priorities when the session starts.” (Participant 32)*

*“I probably talk about the most often with my clients, are both the activity and the sleep.” (Participant 7)*

Overall, clinicians’ perspectives indicated that streaming data use in current practice remains narrow in scope and largely informal. This selective engagement highlights the limited role that streaming data currently plays in routine clinical practice.

### Subtheme 1.3: Patient-Initiated Engagement with Streaming Data Technologies

Clinicians consistently described patient-initiated engagement with streaming data technologies as limited. Most participants reported that patients never (n=10) or rarely (n=16) bring up streaming data technologies during clinical encounters without clinician prompting, indicating that such technology was not a routine component of patient-clinician discussions. Clinicians who described patient-initiated discussions as rare emphasized that engagement occurred sporadically and involved only a minority of patients.

*“They never mention it.” (Participant 19)*

*“I would still say it is very minimal. I’d say, in the last year, it has only been brought to my attention by, maybe, two of my clients. Let’s say less than 10% of clients have brought it up themselves.” (Participant 32)*

Despite overall low levels of engagement, some clinicians observed a gradual increase (n=4) in patient-initiated discussions over the years and only one clinician reporting patients often initiating discussions about streaming data technologies. This suggests that while streaming data technologies remain minimal in most clinical encounters, patient interest may be slowly emerging as streaming data technologies become more widespread and common.

*“I found it’s been definitely increasing in the past couple years.” (Participant 20)*

Together, these perspectives illustrate that patient-initiated use of streaming data in clinical care settings is currently uncommon, but has early signs of increasing.

### Theme 2: Perceived Clinical Value and Usefulness on Streaming Data

Clinicians expressed nuanced perspectives on the perceived clinical value and usefulness of streaming data in health care. While many participants identified specific data types as clinically informative and actionable, others expressed reservations about the utility of other metrics.

Clinicians emphasized that the value of streaming data was highly dependent on context and interpretability, but had an overall positive outlook on its integration into clinical practice.

### Subtheme 2.1: Useful Metrics for Clinicians

Participants identified several types of streaming data that they perceived as clinically useful. Activity and step count data (n=11) was frequently described as valuable for understanding patients’ functioning and engagement.

*“Step counter, so that gives us an accurate number, and then we also have a national benchmarks we can compare that to. So, for example, if someone should walk roughly 7 to 10,000 steps per day, we can see how they’re doing, how active they are compared to that metric. So that helps me, not only having an objective number, but having a benchmark to compare it to.” (Participant 4)*

Sleep data (n=17) was commonly described as clinically meaningful, with clinicians noting their relevance to mental health symptoms.

*“I think especially for people that maybe are coming in, like “I’m feeling…anxious, or I’m having trouble sleeping or, like, I’m maybe feeling more down than usual”. One of the things that I think could be really useful is tracking sleep data.” (Participant 13)*

Cardiovascular metrics (n=17), such as heart rate and heart rate variability, were also viewed as useful.

*“I think most is heart rate and heart rate variability, those two. Heart rate variability because it gives me a nice clean number that we can track the change over time. Like, it went up or down, and it’s one number, it’s not a bunch of metrics, over days. And same with heart rate, and heart rate weighs a little bit more in session, to notice higher points of activation.” (Participant 5)*

Other useful metrics included blood glucose (n=6) and blood oxygen (n=3).

*“Being able to keep an eye on blood glucose levels, that would be useful.” (Participant 15)*

*“Perhaps oxygen saturation levels, because then we could work on breathing strategies and help clients with diaphragmatic breathing.” (Participant 33)*

In addition, some clinicians highlighted the usefulness of medication (n=5), mood, and symptom tracking (n=12), particularly when such data supported adherence monitoring or therapeutic reflection.

*“There’s certain apps, whether it be meditation apps or journaling apps, I guess those are helpful. So then, at least they can log it, and then we can review over, like “what did you write when you were feeling maybe more depressed or not”.” (Participant 19)*

*“Use some type of technology to either link in with an app to say, “yes, I’m taking my meds”, or they would have to scan it at the place that they went to in order to say, “yes I went here, and yes I was observed“.” (Participant 30)*

### Subtheme 2.2: Less Useful Metrics for Clinicians

While some clinicians identified certain streaming data metrics as clinically valuable, others described the same metrics as less useful or difficult to act upon in practice. Differences in perceived usefulness were often tied to challenges related to interpretation, reliability, and alignment with patient experiences. One clinician mentioned that blood oxygen levels would not be a useful streaming data metric. Few participants expressed skepticism about self-reported symptom or stress ratings (n=3), noting limitations in their reliability or interpretive value.

*“I’m torn about the emotional ones where you are, you know, “how do I feel in a given moment”? I feel that depending on their use because it requires a lot of interpretation and sort of reflection as they input that information in, for my clientele, that’s a lot of extra steps.” (Participant 7)*

Sleep metrics (n=4) were also described by several clinicians as difficult to act upon clinically, particularly when the data did not reflect patient reporting.

*“The least actionable is actually sleep, because I find that what it’s tracking versus what client reports aren’t matching. They’re not in alignment. So, they might say, “yeah, I had a really good sleep score, or, it showed that I slept 8 hours, but I woke up feeling exhausted”. And so it just seems a little bit less clinically relevant.” (Participant 5)*

### Subtheme 2.3: Clinical Insights Enabled by Streaming Data

Clinicians described several ways in which streaming data could generate meaningful clinical insights. A key perceived value was the ability of streaming data to provide real-time, longitudinal, and objective metrics that could support more tailored interventions (n=21).

Participants emphasized the importance of observing patterns over time, rather than isolated data points, to better understand their patient’s changes between appointments.

*“What is more important to me is how well have you slept over the two weeks. So, being able to see a pattern in that way, being able to see that, over the last two weeks, what your walking pattern was like, over the last two weeks, what your mood was like.” (Participant 23)*

However, several clinicians stressed that the usefulness of streaming data depended heavily on contextualization (n=6). Without sufficient context, streaming data was viewed as ambiguous and difficult to interpret, limiting their clinical value.

*“I think it’s more just putting it into context to help with clinical decision making. Like, if you just have someone’s heart rate throughout the day, like why did it spike at this time? That could mean anything. That could mean they were working out, that could mean they had to sprint, that could mean a number of different things. So, I think collecting the data could be valuable, but it all just depends on how you contextualize it.” (Participant 13)*

Participants also described streaming data as a way to highlight the connection between physical health indicators and behavioural health conditions (n=4). In particular, clinicians highlighted situations in which streaming data revealed discrepancies between patients’ self-reported experiences and their underlying physiological state, offering additional insight into stress and well-being.

*“Like, you’re telling me you’re fine, but they’re really not internally. But also the building awareness for your physiological state, how much stress you’re actually under versus your self-report of “you’re fine”. There’s a mismatch there, quite frequently, when we’re actually looking at the heart rate, or looking at the heart rate variability.” (Participant 5)*

Streaming data was also described as providing a form of biofeedback for patients (n=7). Clinicians emphasized the potential for such feedback to enhance patients’ somatic awareness and support psychological and therapeutic processes.

*“If they had some sort of wearable device that could give them active feedback around where they’re at in any given moment, it could really help them on a somatic level. Can really help*

*their selves get to know what’s happening in their body because oftentimes that’s our biggest issue we’re coming across, which is people that are really dissociated from their bodies.” (Participant 33)*

Clinicians noted that streaming data could encourage greater patient involvement in their own care by fostering accountability and ownership over health-related behaviors (n=3). Participants described this as a potential mechanism through which streaming data could positively influence engagement and self-management.

*“I think it gives them the opportunity to kind of take accountability and ownership for their own health. That’d probably be the biggest areas where I could see them having a positive impact.” (Participant 20)*

Streaming data was also perceived as particularly valuable for patients with limited access to in-person care (n=3). Clinicians noted that remote monitoring could support ongoing health monitoring for individuals facing geographic, occupational, or logistical barriers to frequent clinical visits.

*“I would say some of the ones would be our patients where we may not have access to care, like ongoing or frequent access to care, whether they live farther out of town or just have other barriers, such as worker life, that prevent them from coming to care.” (Participant 20)*

Despite acknowledged limitations and uncertainties, participants expressed an overall positive outlook toward the integration of streaming data in clinical settings (n=21). Many clinicians emphasized optimism about its future potential and viewed current gaps as opportunities for further development rather than barriers to adoption.

*“So I think that information can be helpful to people, especially when it comes to mental health and sleep. As for opinions, I do actually think this could be a really valuable tool to integrate into the healthcare setting.” (Participant 3)*

*“I want to encourage it. And us not being there today or not having the most appropriate data is not a disincentive. But it’s an incentive centered towards doing better, finding alternative ways of doing things, developing better sensors, better algorithms, better software, all that kind of stuff.” (Participant 23)*

Overall, clinicians described streaming data as a way of providing additional clinical insight, from identifying longitudinal patterns and physiological-behavioural connections to supporting patient engagement.

### Theme 3: Usability, Accessibility, and User Support

Clinicians emphasized that the usability and accessibility of streaming data technologies were central to whether such data could be meaningfully incorporated into current clinical workflow systems. Participants highlighted that both how data are presented to clinicians and the integration into current electronic medical record systems strongly shaped perceived feasibility and usefulness.

### Subtheme 3.1: Understandability and System-Level Accessibility of Streaming Data

Clinicians emphasized that streaming data would be most usable when presented in a clear, easily understandable manner (n=14). Participants described the need for uniform data formats and consolidated displays that would allow clinicians to quickly understand key information without navigating multiple devices, platforms, or granular time points. Participants further highlighted the importance of integrated, summative views of streaming data that link multiple data types and emphasize clinically meaningful information. Rather than raw data streams, clinicians described a preference for data summaries that highlight key trends or discussion points.

*“It can be all in one. So, the more data points, the more data, the type of data, will be much more comprehensive, and link the information. And if it’s summative, and post-analytic data, and highlights key points, highlights key information, and provides a statistic, or meaningful discussion point, that would be really important.” (Participant 9)*

*“Making sure that it’s uniform, one center has all of these different streaming units of measures, so that we don’t have to learn so many different devices. And then having a summary in that streaming application, just so that we don’t have to go through, day by day, or hour by hour, or minute by minute kind of thing.” (Participant 22)*

In addition to clear data presentation, clinicians highlighted the importance of system-level accessibility through seamless integration with existing electronic medical record (EMR) systems (n=21). Participants described the need for streaming data to flow directly into patient charts, allowing clinicians to access information efficiently and in advance of clinical encounters.

*“So, we need the data to come into the patient’s chart, so we use a unified health system electronic medical record within our Health Authority, and it’d be nice if whatever we chose and we wanted to do can integrate it currently. It would be better if it could be all seamlessly integrated, so perhaps the patient can have an app that delivers that information to us.” (Participant 4)*

*“So, I think having an easy-to-integrate system, so that patients are able to upload their data and sort of real-time or in short period of time be uploaded onto the EMR so that I’m able to access those records before a patient shows up to the clinic.” (Participant 27)*

These findings indicate that clinicians view clear data presentation and seamless EMR integration as foundational requirements for the practical use of streaming data in routine clinical care.

### Subtheme 3.2: Educational Supports and Learning Clinical Applicability

Clinicians emphasized the need for streaming data technologies to be easy to use and supported by appropriate educational resources for both clinicians and patients. Clinicians noted that training was necessary to raise awareness of available tools and to support them in introducing these technologies appropriately during clinical encounters (n=8).

*“I think, first of all, having some teaching about it, telling us what’s available, how to go about presenting it to patients.” (Participant 15)*

Participants also highlighted the importance of clear, accessible education resources for patients (n=12), particularly at the point of device adoption. Clinicians suggested that standardized onboarding, such as modules or information sessions, could support patient understanding of device use and data sharing, while also clarifying how streaming data can contribute to their clinical assessment and care planning.

*“I think that could be managed through certain modules or information sessions when they first purchase these devices or use these devices. And then they can have a straightforward walkthrough in terms of how to use the device, and then upload the data, or how it could be used for the physician and benefits their overall assessment and plan.” (Participant 27)*

Clinicians also emphasized the need for ongoing user support to minimize disruptions to clinical workflows. Participants described the value of having designated support personnel or representatives to assist patients with technology-related questions, reducing the burden on clinicians’ time.

*“If there was a representative that I could direct someone to, for how to use a certain technology then that would be really helpful. Because again, then it would just not take time away from my own workflow.” (Participant 1)*

In addition to operational support, clinicians highlighted the need to learn how streaming data can apply in clinical contexts (n=9). Participants described a need for support in understanding what different data patterns might signify and how such information could inform clinical decision-making.

*“So knowing what to look at, and kind of when to look. And then on top of that learning it would be like learning a new clinical sign or symptoms, like understanding what it means. So having some assistance in understanding, like, if the steps are trending down, and their heart rate variability is also trending down, like, what does that mean? So just some interpretation would be helpful.” (Participant 12)*

Overall, clinicians described that without education, user support, and guidance on clinical applicability, streaming data technologies would remain difficult to integrate into routine practice despite their potential value.

### Theme 4: Feasibility, System Barriers, and Workflow Challenges

Clinicians described multiple feasibility challenges that limited the integration of patient-generated streaming data into routine clinical care settings. These challenges spanned across system-level barriers, workflow constraints, and broader organizational and patient-level considerations. Overall, participants emphasized that without addressing these barriers, streaming data technologies would remain difficult to implement at scale.

### Subtheme 4.1: System and Workflow Barriers to Integrating Streaming Data

Clinicians identified challenges related to integrating patient-generated streaming data into existing electronic medical record systems. Participants described difficulties with bringing new technologies into current clinical systems (n=5).

*“Our EMRs themselves are very rigid and they’re not supportive of new technology.” (Participant 23)*

Time constraints within clinical practice were also cited as a major barrier to streaming data use (n=4). Clinicians emphasized that reviewing and interpreting additional data streams was difficult to accommodate within already limited appointment times.

*“We have a very limited amount of time with the patients, so I can foresee that being difficult.” (Participant 22)*

*“I mean, first and foremost, the time involved to integrate something could be a challenge, at least for my practice, would be very difficult.” (Participant 33)*

In addition to time pressures, participants described the volume of streaming data would be overwhelming (n=5). Managing and sorting through large amounts of continuously generated data was perceived as challenging and potentially unmanageable in clinical settings.

*“I think just the overwhelming amount of information, I can foresee that being a difficult aspect.” (Participant 29)*

Together, these perspectives highlight that feasibility concerns related to system integration, time limitations, and data volume pose significant barriers to incorporating streaming data into routine clinical workflows.

### Subtheme 4.2: Organizational, Patient, and Resource Considerations

Participants emphasized that successful implementation of streaming data technologies would require broader buy-in from the entire healthcare team (n=11). Clinicians noted that inconsistent adoption across all members of the healthcare team will limit feasibility and create fragmentation in care.

*“And people, not only a patient, but the caregiver, care team will work on those. So this would need leadership, from a health region, doctor training system, and also public media.” (Participant 6)*

*“I think just willingness from everyone. Of course, the clinicians have to be willing to incorporate into their practice, but if there’s a manager that’s not willing to, then it’s not gonna have the buy-in.” (Participant 13)*

Participants also described that streaming data technologies would not be feasible for all patients (n=18). Use was often limited to certain patient groups, particularly when cost or access to technology was a concern.

*“If the patient has the technology. Some of my patients don’t even have a smartphone, so if they’re struggling with houselessness and things like that, they might not have even a phone to do it. So then we might need to have that technology in the clinic, for them to input some data.” (Participant 4)*

*“I have a very elderly practice, number one. And a lot of the elderly patients do not have technology access. Most of my patients are 55, 65 plus to 104 years of age. So, a lot of the elderly with mental health issues will not have technology. That’s problem one. Problem number two, a lot of patients with major mental health issues cannot afford technology.” (Participant 16)*

Patient compliance and technological competency were further described as essential for streaming data integration (n=8). Clinicians emphasized that effective use depended on patients’ ability to manage and reliably engage with the technology.

*“It comes down to the individual user, their level of competency and awareness and understanding of what they are.” (Participant 23)*

*“The challenge is just around operator error. I mean, you try to give as much instructions and details and people interpret things differently than they’re supposed to be. So it often can, either inaccuracies or loss of data, or no data, and then that can run into frustrations with the patient as well.” (Participant 24)*

Finally, clinicians highlighted the importance of considering financial costs associated with integrating streaming data technologies into clinical settings (n=8). Participants raised questions about the cost to the clinic and how this might impact adoption.

*“Cost is what can be considered, right? Because let’s just say you have a very basic electronic medical record, and then you want to integrate more data and an additional process, some of these companies that make these electronic medical records, they’ll probably end up charging you for it.” (Participant 18)*

*“I think money’s gonna be a big one. Our practice is pretty big, like, we have a lot of staff, we have a lot of resources. I think if we wanted to implement something, it’s maybe can be a little bit easier than a smaller clinic.” (Participant 26)*

Collectively, these perspectives illustrate that feasibility of streaming data integration extends beyond individual clinician interest, encompassing organizational alignment, patient capability, and financial considerations that must be addressed to support sustainable and widespread implementation.

### Theme 5: Data Quality, Reliability, and Validity

Clinicians expressed differing views about the quality, reliability, and validity of streaming data technologies, voicing concerns about some metrics while expressing confidence in others.

### Subtheme 5.1: Variability in Reliability and Validity of Streaming Data

Most clinicians expressed reservations about the reliability and validity of streaming data and emphasized that further consideration and evaluation are required before these data can be used to guide clinical decision-making or tailor care plans (n=25). Participants described concerns about understanding the strengths and pitfalls of different devices, drawing parallels to traditional diagnostic testing and emphasizing the absence of consensus regarding which technologies were sufficiently reliable.

*“And then another challenge-issue barrier would be the validity of the data. So, understanding the pitfalls in the data the same way that we do about a single test, like the positive predictive value, negative predictive value.” (Participant 12)*

*“The information has to be taken with a grain of salt before using it for clinical purposes or initiating treatment or diagnosis.” (Participant 18)*

In contrast, some clinicians (n=7) expressed confidence in the reliability of streaming data, particularly when data were collected continuously or longitudinally. Participants described greater trust in subjective and longitudinal measurements in mental health patients, and noted increasing confidence in the overall quality of collected data. Confidence was especially evident for objective measures with established clinical validation, which are described in greater detail in Subtheme 5.2.

*“I would trust it because I think with mental health, a lot of it is, when you ask about symptoms, they change very quickly for a patient. So, if I had access to data that was taken over several periods of time, or continuous time period, then I would actually trust it more than what the patient is saying to me right then and there. If it’s subjective, then again, I would trust it, because again, the patient’s gonna tell me, anyways, what they think.” (Participant 1)*

Together, these perspectives illustrate that clinicians’ confidence in streaming data reliability is not uniform, reflecting both ongoing skepticism and growing trust as technologies mature.

### Subtheme 5.2: Confidence in Specific Streaming Data Metrics

Clinicians described confidence in the reliability and validity of specific streaming data metrics, with greater trust placed in measures that are objective, clinically familiar, and supported by established validation practices. Blood glucose monitoring technologies (n=1) were explicitly mentioned as reliable, especially in the context of diabetes care.

*“Patients with diabetes, I trust the DexCom, and I trust the freestyle Libra. Those are fine.” (Participant 18)*

Heart rate monitoring data (n=2) were similarly described as relatively reliable, particularly when the measurements were objective and numerical, and when accuracy had already been established in real-world settings.

*“Heart rate I might trust that a bit more. So, if it is objective and it is numerical, it has to actually be accurate in the community already.” (Participant 1)*

Blood pressure monitoring technologies (n=2) were also described as trustworthy, particularly when combined with clinic calibration and verification processes to ensure accuracy.

*“So, fairly confident in the accuracy of those, as well as our Holter monitors, and we do 24-hour blood pressure monitors as well. But again, that’s because we do a lot of our own calibration and verification with regards to the values.” (Participant 24)*

### Theme 6: Privacy and Regulation Considerations

Clinicians discussed privacy and regulatory considerations as important factors shaping the acceptability and implementation of patient-generated streaming data in clinical care. While perspectives varied, participants emphasized the need to balance potential clinical benefits with appropriate safeguards related to data privacy, consent, storage, and regulatory oversight.

### Subtheme 6.1: Privacy, Consent, and Secure Handling of Streaming Data

Some clinicians reported minimal concerns regarding privacy or data sharing when streaming data consisted of relatively low-risk physiological or behavioral measures (n=3). These participants differentiated between streaming data types, noting greater comfort with commonly collected metrics such as movement or heart rate.

*“I actually have very few privacy concerns right now. If we were collecting different data, EEG data particularly, I’d have many more concerns. But the privacy data of what my pulse is and what my movement is, and what my activity is, isn’t the great concern.” (Participant 17)*

In contrast, more clinicians expressed ongoing concerns related to privacy, consent, and the potential misuse of patient-generated streaming data (n=13). Participants highlighted risks associated with digital data collection. Concerns were also raised about data storage and access particularly when third-party platforms or electronic systems were involved.

*“Privacy is always, always a concern, right? I guess nowadays everything’s digital, right? So, information’s readily accessible, but also there’s always risk of it being stolen or used for other purposes.” (Participant 18)*

*“How are we protecting, how we’re using this information? And who has access? And I guess it goes also over to, if we are using an online platform or form or some sort of electronic health record, where is the data being stored? I think that’s one of the major concerns.” (Participant 33)*

Clinicians emphasized the importance of transparency and informed consent, both for patients and clinicians, regarding what data are collected, how they are used, and with whom they are shared (n=13). Participants also highlighted a desire for greater clarity around how consumer-generated data are handled by external companies before being transmitted to clinicians.

*“And storage, making sure that clients have informed consent about what they’re sharing and how it will be used. And do they really understand? Wanting that informed consent so that they understand what we’re collecting, why we’re collecting it, and how we’re going to use it. And then how it will be stored and destroyed at whatever point in time. And whom it will be shared with.” (Participant 3)*

*“I think that’s probably going to be on the consumer side of things. If they’re going to be using these technologies on their own accord, they probably would have some kind of, maybe sign, a terms and conditions or agreement with that external company. But then I think I would also kind of want to know how that data is being stored or manipulated before it gets sent to me.” (Participant 26)*

Secure and de-identified data handling methods were consistently described as essential for mitigating privacy risks (n=10). Clinicians emphasized the need for encryption, anonymization, and adherence to existing privacy protocols when managing streaming data.

*“If we could find a way to encrypt data, so just in case it gets released, no one could identify you. Like, maybe if there was some sort of protocol that we could follow that would ensure that those private, a way to identify a patient, is not used in the data collection.” (Participant 2)*

*“Getting that data and getting it in a private, secure way that fits with all of their very, very, very long policy documents around privacy and medical information and that sort of stuff.” (Participant 10)*

Together, these perspectives underscore that clinicians’ acceptance of streaming data is closely tied to assurances around privacy, consent, and secure data handling, setting the foundation for regulatory and governance considerations.

### Subtheme 6.2: Regulatory Guidelines and Ethical Use of Streaming Data

Clinicians emphasized that streaming data technologies should be vetted and approved by appropriate authorities before being integrated into clinical care (n=12). Participants described regulatory approval as a way to establish trust, standardization, and legitimacy. Compliance with provincial and federal regulations governing data privacy and storage was also identified as a prerequisite for clinician comfort and adoption.

“This product should be utilized, verified, and approved by the health authority, so this way people know this is a standard of care or standard tool for care or diagnosis.” (Participant 6)

*“As long as they’re doing their proper due diligence to ensure that they’re complying with provincial and federal Canadian laws for data storage and encryption and privacy, I think that’s probably the first thing I’d be looking for, to ensure that I would be comfortable using it. So definitely Freedom of Information and Protection of Privacy Act and Personal Information Protection Act compliant.” (Participant 33)*

Beyond regulatory compliance, clinicians emphasized that streaming data should be used strictly to support patient care (n=5). Participants expressed concerns about data being accessed or leveraged for non-clinical purposes. Some clinicians raised specific concerns about potential downstream use of streaming data by third parties, such as insurers, and the possible negative consequences for patients.

*“I just want to make sure that information goes to the EMR and only for patient record and not for any other purposes.” (Participant 18)*

*“If I wanted this information shared, I’d want to share it for benefits. My concern comes that when this information is shared, the first people that are usually able to get a hold of it or utilize it, are actually going to be insurance companies. And similar entities that will either use it to make your life more difficult, limits your freedoms, or increase your premiums, or limits your ability to get disability coverage, or something like that.” (Participant 23)*

Collectively, these perspectives highlight that clinicians view privacy protection, regulatory oversight, and ethical use as foundational requirements for the responsible integration of streaming data into clinical mental health care.

## Discussion

This study provides an in-depth examination of clinicians’ perspectives on the integration of patient-generated streaming data into routine mental health care. Across interviews, clinicians described streaming data as holding substantial conceptual promise for enhancing clinical monitoring, particularly by capturing longitudinal fluctuations in behavioral and physiological functioning that occur between clinical encounters. Participants emphasized that the value of streaming data is shaped by multiple interrelated factors, including the interpretability of data outputs, alignment with clinical workflows, organizational readiness to support implementation, confidence in data reliability, and the presence of clear governance and regulatory safeguards.

Collectively, these findings indicate that clinicians are not resistant to streaming data technologies, but rather take a cautious, context-dependent approach to their use. Importantly, successful translation into routine care appears to hinge less on the availability of data and more on how well these technologies align with the practical, organizational, and regulatory realities of clinical practice.

Clinicians articulated a clear preference for streaming metrics that map onto established clinical constructs and an emerging evidence base. The domains most frequently identified as relevant, such as sleep, physical activity, and heart rate, mirror those commonly examined in wearable-based mental health research and have been linked to symptom severity, illness trajectories, and functional outcomes across conditions [38,39]. This alignment suggests that perceived usefulness is driven less by technological novelty and more by whether a metric is recognizable, interpretable, and clinically actionable [40].Thus, streaming measures that objectively reflect patterns already discussed in routine assessment may be easier to integrate, as they are understood as extensions of familiar clinical reasoning rather than wholly new forms of information [41]. Clinicians also differentiate between “useful” data and simply “more” data, emphasizing that high-volume or ambiguous information has limited value unless it can inform decisions or prompt meaningful clinical action [26,42,43].

Building on this, clinicians emphasized that the central contribution of streaming data is not merely the availability of additional metrics, but the potential to transform how mental health is monitored and interpreted over time. Clinicians described streaming data as enabling a more dynamic and individualized understanding of symptom trajectories by capturing temporal fluctuations in behavioural and physiological functioning that can be missed by traditional episodic, retrospective assessment. Emerging evidence suggests that continuous monitoring of sleep patterns, activity variability, and physiological stress indicators can provide valuable insight into early symptom changes and relapse risk across mood and anxiety disorders, supporting the development of more proactive and personalized models of care [43,44].

Clinicians further noted that objective behavioral and physiological signals could add context to patient-reported experiences, addressing recognized limitations of recall-based reporting and potentially strengthening diagnostic clarity and treatment monitoring [15]. Clinicians also perceived potential therapeutic value of streaming data by supporting shared decision-making, enhancing patient engagement, and promoting self-management when trends are visualized in clinically meaningful ways [45,46]. Across these anticipated benefits, clinicians repeatedly returned to one condition: clinical impact depends on translating complex data streams into synthesized, contextualized outputs that can be interpreted and acted upon within routine care [26,47,48].

Implementation barriers were described as one of the primary determinants of whether streaming data could move from bench side to bedside. Clinicians highlighted that digital health tools often encounter adoption challenges when technological capability outpaces clinical infrastructure, particularly interoperability with EMRs and the realities of time-limited appointments [27,49,50]. Disconnected platforms, manual data, or poor data access can disrupt workflows and increase administrative burden, ultimately undermining engagement even when the underlying metrics are clinically relevant [51,52]. These challenges reflect that digital tools can disrupt established clinical workflows and contribute to increased workload and technostress when integration strategies do not align with real-world practice demands [53,54]. Many clinicians emphasized the need for streaming data to be synthesized into concise, clinically meaningful summaries. Large volumes of raw metrics were viewed as difficult to interpret and impractical to act on without intuitive visualizations and clear contextualization; thus, reinforcing that wearable-derived data only become clinically useful when presented in formats that foreground actionable insights [40]. Clinicians also highlighted organizational constraints, including time pressures, staffing limitations, and the lack of institutional support for training or technical troubleshooting. Literature emphasizes that organizational readiness, including leadership endorsement, resource allocation, and workflow adaptation, is essential for adopting new technologies in health care [55,56]. Collectively, these accounts suggest that usability, interoperability, and organizational support are not peripheral considerations; they are prerequisites for routine clinical use.

Clinician confidence represents another key determinant of streaming data integration into mental health care. Evidence suggests that adoption of patient-generated health data is strongly influenced by the availability of validated metrics, clear interpretable frameworks, and demonstrated clinical utility [57]. In the absence of these supports, patient-generated streaming data can introduce uncertainty and increase cognitive burden, limiting their integration into clinical decision-making despite technological advancement [25,26,58]. Clinicians described hesitancy to rely on streaming data when they could not confidently determine what a given pattern meant in context or how much weight it should carry relative to patient report and clinical presentation. As a result, even when streaming data were viewed as potentially informative, limited trust in their validity and interpretability reduced clinicians’ willingness to incorporate them into care planning.

Regulatory and governance considerations represent a fundamental requirement for the safe and sustainable integration of streaming data into mental health care. Existing digital health literature highlights that trust in patient-generated health data is strongly influenced by transparent governance structures addressing privacy protection, data ownership, and responsible data use [59,60]. Continuous monitoring introduces additional ethical and legal complexity by producing sensitive behavioral and physiological information outside conventional clinical documentation boundaries and often involving third-party data handling [61–63]. Concerns regarding data security, third-party data handling, and potential downstream use of patient-generated information have been widely identified as barriers to clinician and patient acceptance of digital monitoring technologies [46,64,65]. Uncertainty about professional liability and accountability when streaming data signal potential clinical risk remains a major unresolved implementation barrier, underscoring the need for clear regulatory frameworks and institutional policies that define monitoring expectations, establish responsibility, and support ethical use in ways that build clinician trust [65,66]. These findings reinforce the need for governance models that define data stewardship, consent processes, monitoring responsibilities, and legal accountability in order to support the responsible integration of streaming data into routine clinical practice.

Taken together, this study demonstrates that clinicians view streaming data as a promising extension of mental health monitoring, particularly for enhancing longitudinal assessment and supporting personalized care. However, the successful translation of streaming data from technological innovation to routine clinical use is shaped by the interplay between clinical interpretability, workflow integration, organizational readiness, clinician confidence, and regulatory clarity. Importantly, clinicians occupy a central role in determining whether patient-generated data are incorporated into clinical decision-making, highlighting the importance of clinician engagement in the development and implementation of streaming data technologies. Strategies such as co-design with clinicians, investment in interoperable infrastructure, and establishment of clear governance and regulatory safeguards may help move streaming data from innovation to sustainable practice. Without attention to these contextual conditions, streaming data risk remaining underutilized despite their potential to enhance monitoring, engagement, and continuity of care.

## Limitations

This study should be interpreted with the following limitations in mind. First, the findings reflect the perspectives of a purposively recruited sample of clinicians who were willing to participate in interviews about digital and streaming data technologies. Although the sample included a diverse mix of family physicians, psychiatrists, and psychologists, clinicians with stronger opinions or greater interest in digital health may have been more likely to participate. Thus, the views captured may not fully represent those of clinicians who are less engaged with, or more resistant to, digital health and streaming data technologies, potentially influencing the range of perspectives identified. Future research could address this limitation by deliberately sampling clinicians with low digital adoption, limited exposure to streaming data, or explicit skepticism toward digital tools. Additionally, this study relied on self-reported perceptions and experiences rather than direct observation of clinical workflows or real-world implementation of streaming data systems. As such, clinicians’ perspectives may be influenced by recall bias. Future studies incorporating observational methods, workflow analyses, or implementation studies could provide more insight into how streaming data is functionally integrated, interpreted, and acted upon in routine clinical practice.

Moreover, while reflexive thematic analysis allowed for rich, in-depth exploration of clinician perspectives, the findings are not intended to be statistically generalizable. The study prioritizes conceptual and experiential insights over prevalence estimates, and therefore the frequency of codes or themes are not indicative of their relative importance across all clinical contexts. Future research could build on these qualitative findings through mixed-methods or quantitative studies designed to assess the prevalence and variability of identified themes across larger and more diverse clinician populations.

Finally, although analytic rigor was supported through team-based coding, collaborative review, and iterative theme refinement informed by Braun and Clarke’s reflexive thematic analysis, qualitative analysis is inherently interpretive and researcher-driven. The themes presented therefore reflect one plausible interpretation of the data and are shaped by the theoretical orientations, disciplinary backgrounds, and positionality of the research team [36,67]. While regular team discussions were used to reflect on potential analytic influences, more formal reflexive practices, such as systematic documentation of researcher positionality, could have further enhanced transparency and critical appraisal of the analytic process [68–70]. Even with the use of multiple coders and iterative discussion to strengthen analytic credibility, these strategies do not fully eliminate subjectivity or ensure that all perspectives were equally represented.

## Additional Information

### Data Availability

The original contributions of this study are included in the article.

### Institutional Review Board Statement

This study was approved on September 26th, 2025, by the Research Ethics Board of the University of Alberta (Pro00155136).

